# Nitrous Oxide Alters Functional Connectivity in Medial Limbic Structures in Treatment-Resistant Major Depression

**DOI:** 10.1101/2024.08.12.24311729

**Authors:** Charles R. Conway, Ben Julian A. Palanca, Thomas Zeffiro, Britt M. Gott, Frank Brown, Victoria de Leon, Linda Barnes, Thomas Nguyen, Willa Xiong, Christina N. Lessov-Schlaggar, Gemma Espejo, Steven Mennerick, Charles F. Zorumski, Peter Nagele

**Author notes:** <u>Corresponding Author:</u> Charles R. Conway, MD Professor of Psychiatry, Departments of Psychiatry and Radiology 660 South Euclid, Campus Box 8134.

## Abstract

While nitrous oxide (N_2_O) has demonstrated antidepressant properties in treatment-resistant major depression (TRD), little is known about neural mechanisms mediating these effects. Employing serial resting-state functional magnetic resonance imaging (rs-fMRI), we compared spatiotemporal effects of inhaled N_2_O on brain functional connectivity in TRD patients (n=14) and non-depressed healthy controls (n=16, CNTL). Participants received sequential, one-hour inhalations of either 50% N_2_O/oxygen or air/oxygen (placebo), with sessions separated by at least one month in random cross-over order. BOLD-contrast rs-fMRI scans were acquired at three time points: pre-inhalation, 2 hours post-inhalation, and 24 hours post-inhalation. For the rs-fMRI functional connectivity analyses, five *a priori* seeds in medial limbic structures targeted cortical networks implicated in major depression – the salience, anterior and posterior default mode, reward, and cingulo-opercular networks – and a nexus in the dorsal paracingulate region previously identified in MDD (“dorsal nexus”). Depression, dissociation, and psychosis assessments were made before and after inhalations. In TRD patients, functional connectivity was reduced in all seeded networks and the voxel-wise global analysis after N_2_O exposure. N_2_O progressively *decreased* connectivity in patients with TRD but *increased* connectivity in healthy controls. In TRD patients, each seeded network demonstrated post-inhalation functional connectivity reductions in the dorsal paracingulate gyrus (“dorsal nexus”). This study further elucidates neural mechanisms underlying the antidepressant properties of N_2_O, supporting the notion that N_2_O specifically alters mood-associated brain regions in the depressed brain state by reducing functional connectivity within these brain networks.

The trial was registered at ClinicalTrials.gov (NCT02994433).

**One sentence summary:** Nitrous oxide reduces long-range functional brain connectivity in treatment-resistant major depression, which may underlie its antidepressant action.

## INTRODUCTION

Nitrous oxide (N_2_O) is an N-methyl-D-aspartate receptor (NMDAR) antagonist, known colloquially as “laughing gas.” N_2_O has demonstrated promising therapeutic results for several psychiatric disorders, including TRD (1–3), non-resistant major depressive disorder (MDD; 4), bipolar depression (5), post-traumatic stress disorder (PTSD; 6), and suicidality (7).

Two double-blind, placebo-controlled trials have demonstrated rapid and sustained effects in TRD with a single one-hour inhalation of 50% or 25% inhaled N_2_O (1, 2). N_2_O is well-tolerated as a treatment for MDD and TRD, with its antidepressant effects demonstrating rapid onset and typically lasting at least 1-2 weeks (1–4).

Similar to ketamine, N_2_O’s antidepressant effects are both rapid and sustained. Exposure effects are observed for days to weeks after a single dose, suggesting that N_2_O produces lasting CNS changes that persist beyond its acute brain interactions. These lasting effects may be mediated via neural plasticity within brain networks (8, 9) and changes in correlated neural activity, i.e., functional connectivity. Our group reported sustained alterations in functional connectivity in the visual and dorsal attention networks after N_2_O exposure in non-depressed healthy subjects (10). Nevertheless, little is known regarding how N_2_O alters functional brain networks to confer antidepressant effects.

Although neural mechanisms underlying depression remain incompletely characterized, current major depressive disorder/treatment-resistant depression (MDD/TRD) models posit involvement of five overlapping brain networks (11, 12). These include the reward network (RN), the default mode network (DMN), the cingulo-opercular network (CON), an executive control circuit (13), and the salience network (SN) (12, 14–17). Previous imaging studies have demonstrated spatially convergent depression-related differences involving three of these networks (CON, DMN, and RN) in the dorsal paracingulate gyrus, referred to as the “dorsal nexus”, a region that shows markedly higher connectivity in MDD compared to non-depressed controls (18). The goal of this study was to characterize changes in functional connectivity and depressive symptoms resulting from N_2_O exposure in patients with severe TRD and CNTL participants in these brain networks.

## Results

### Demographics and Data Quality

**Supplemental Figure 1** (CONSORT diagram) summarizes participant screening; further details of participant screening are found in the Results section of the supplement. The final sample of 14 TRD participants is summarized in **Table 1S** and included 10 females, with a mean age of 42.5 years, a mean of 21 years of lifetime MDD, and a mean of 7.6 lifetime failed antidepressant trials. The final CNTL sample is summarized in **Table 2S** and consisted of 16 participants, including 9 females, and a mean age of 36.4 years. The TRD cohort target sample size of 20 TRD and 20 CNTL was prespecified in the CT.gov registry but was not met due to early termination of the study during the COVID-19 pandemic.

### Summary of rs-MRI Functional Connectivity Analyses

For all seeded and global functional connectivity findings, a consistent pattern emerged: N_2_O exposure resulted in *decreased* connectivity in patients with TRD, whereas in non-depressed controls, N_2_O exposure resulted in *increased* connectivity. Supplementary **Table 3S** summarizes differences in network connectivity observed in TRD and CNTL. **Figure 2 middle panel and bottom panels** demonstrate the mean combined pre-post average network changes across all 5 seeds following N_2_O and air (placebo) exposure, respectively.

**Figure 1.**
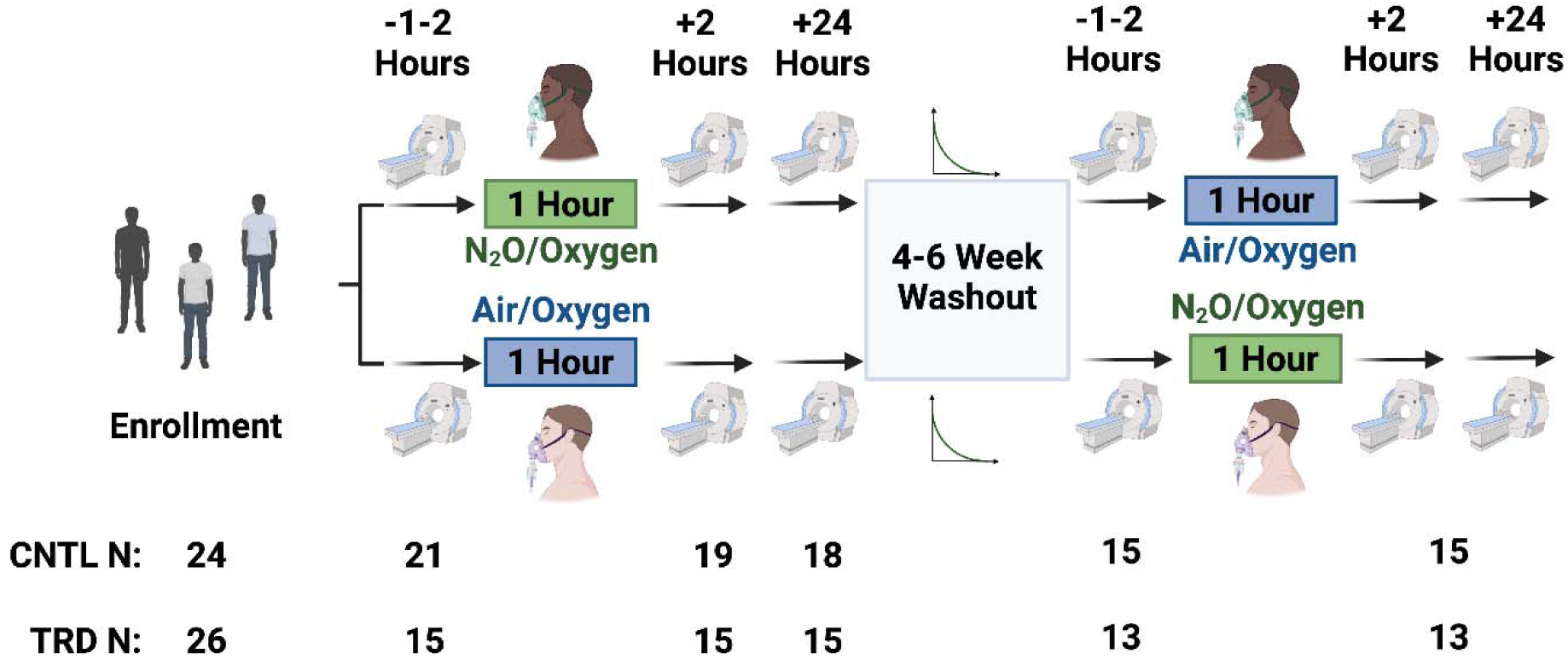
Study design summary. All participants, both treatment-resistant depression (TRD; N=14) and non-depressed controls (CNTL; N=16) were randomized to receive either 50% inhaled nitrous oxide (N_2_O) or placebo (air/50% oxygen mixtures). One to two hours pre-inhalation, participants underwent a 30-minute pre-exposure fMRI scan, followed by a one-hour inhalation session, followed by sequential thirty-minute fMRI scans at 2- and 24-hours post-exposure. This process was then repeated following a 4–6 week washout period. Data from one TRD and two CNTL participants who withdrew after N_2_O inhalation and imaging was included in the analyses.

**Figure 2.**
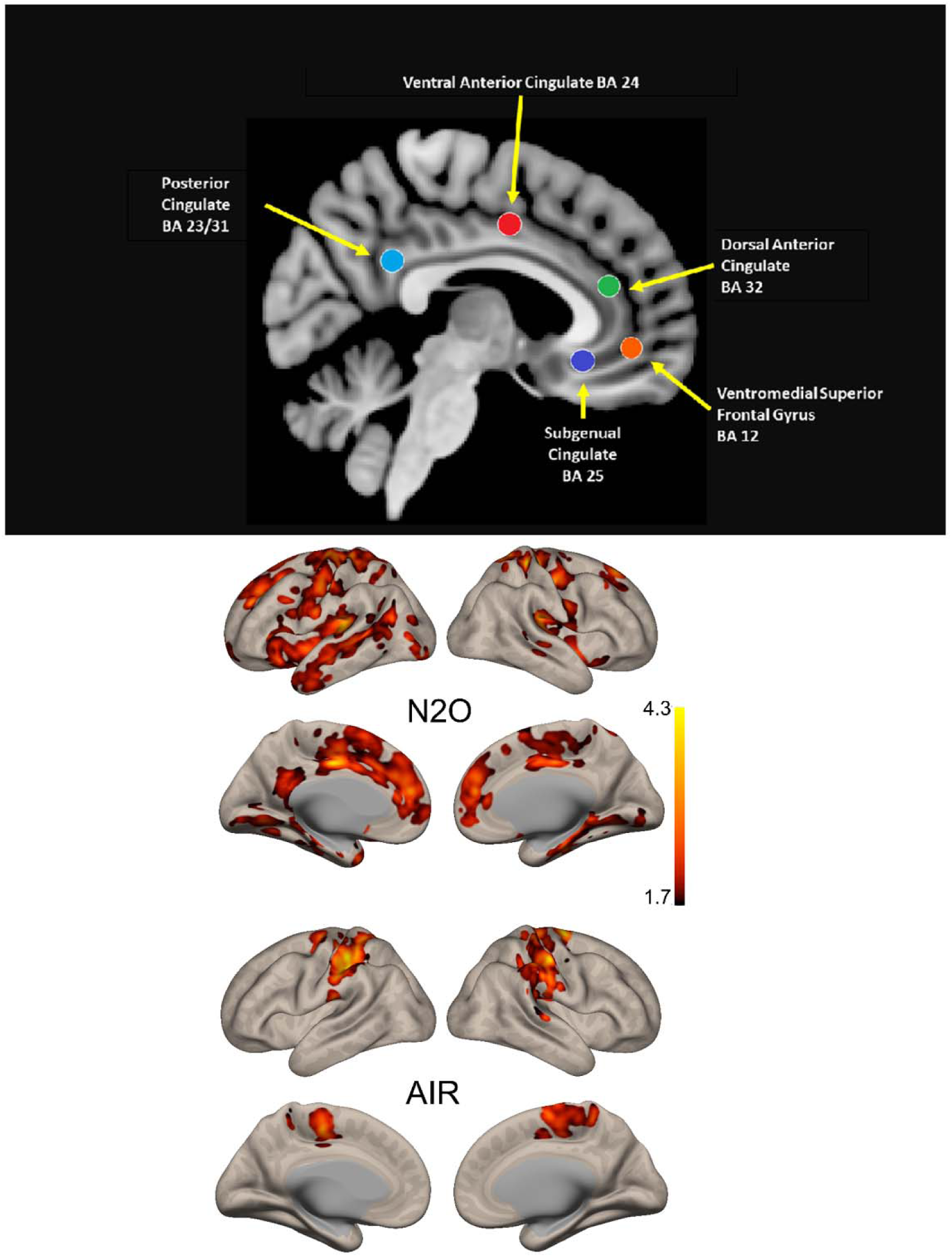
Seeds (top panel) and Changes in Functional Connectivity Observed Before and After N_2_O (middle panel) and AIR (bottom panel). <u>Top Panel</u>: *A priori* seeds were selected to compare pre-post exposure to N_2_O and placebo. These five seeds targeting midline limbic structures probed networks known to be involved in mood regulation: the reward network (Brodmann’s area BA 25), the cingulo-opercular executive network (midline dorsal cingulate, BA 24,red), the salience network (dorsal anterior cingulate, BA 32, green), and the anterior and posterior nodes of the default mode network (ventromedial prefrontal cortex, BA 12, orange, and posterior cingulate, BA31, cyan, respectively). <u>Middle</u> <u>Panel</u>: Changes in functional connectivity averaged across all 5 seeds comparing pre- to 2 and 24 hours post-N_2_O exposure. There are notable changes in functional connectivity observed in the anterior and posterior portions of the cingulate cortex, hippocampal and parahippocampal regions, and insular cortex. <u>Lower Panel</u>: Changes in functional connectivity averaged across all 5 seeds comparing pre- to 2- and 24-hours post-placebo exposure. We observed markedly different spatial changes in functional connectivity, with changes limited to primarily parietal cortex and no involvement of the cingulate, hippocampus, insula or dorsal paracingulate gyrus.

These findings demonstrate, as detailed below, that over the course of 24 hours, inhalation of N_2_O is consistently associated with *reductions* in functional connectivity in TRD and *increases* in functional connectivity in CNTL participants. Further, N_2_O exposure selectively affects functional connectivity with multiple regions known to be associated with mood regulation, including the dorsal paracingulate gyrus (“dorsal nexus”) previously reported to have higher functional connectivity in MDD (16).

### Individual Seed Findings

We observed changes in functional connectivity for each of the five seed -> voxel maps separately. Pre-post exposure regional changes in functional connectivity for the combined group (CNTL + TRD) were observed over time for N_2_O (**Figure 3**) and placebo (**Figure 4**) and show distinct patterns.

**Figure 3.**
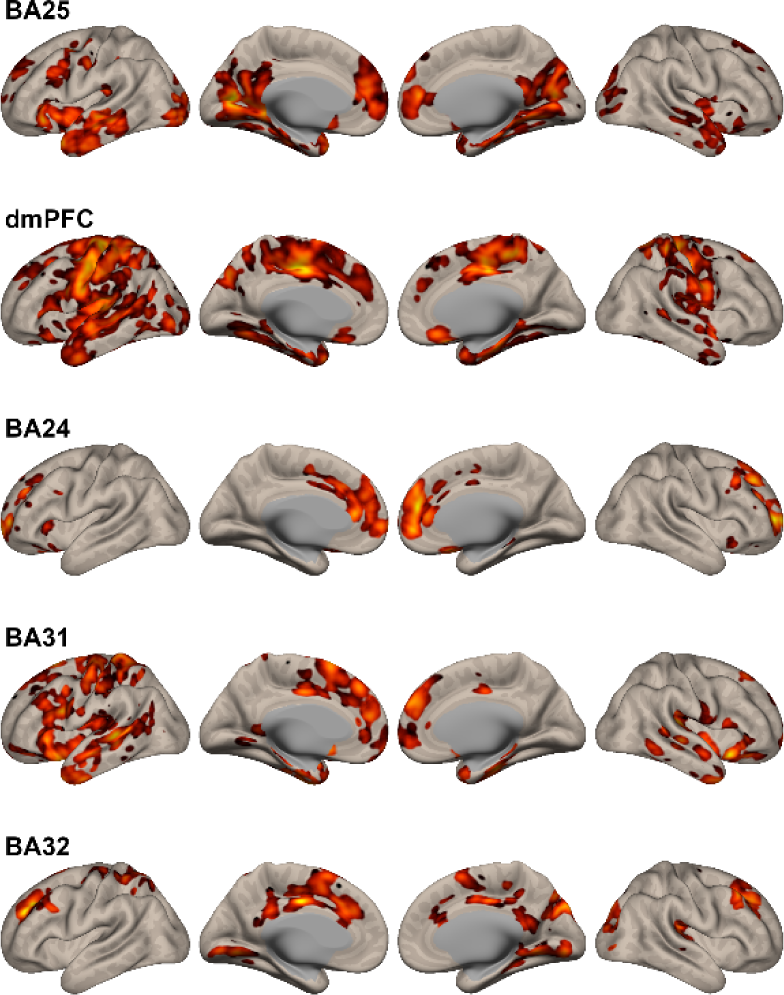
Changes in Functional Connectivity Observed After N_2_O Exposure. Details of the regional changes observed for each seed are described in the text and Table 3S. In general, changes in functional connectivity in regions associated with mood regulation were observed, including the medial prefrontal cortex, anterior cingulate, paracingulate cortex, hippocampus, and parahippocampus. Additionally, all seeds demonstrated changes in functional connectivity in the dorsal paracingulate gyrus, previously identified as having increased resting-state connectivity in MDD as the “dorsal nexus” (Sheline et al., 2009).

**Figure 4.**
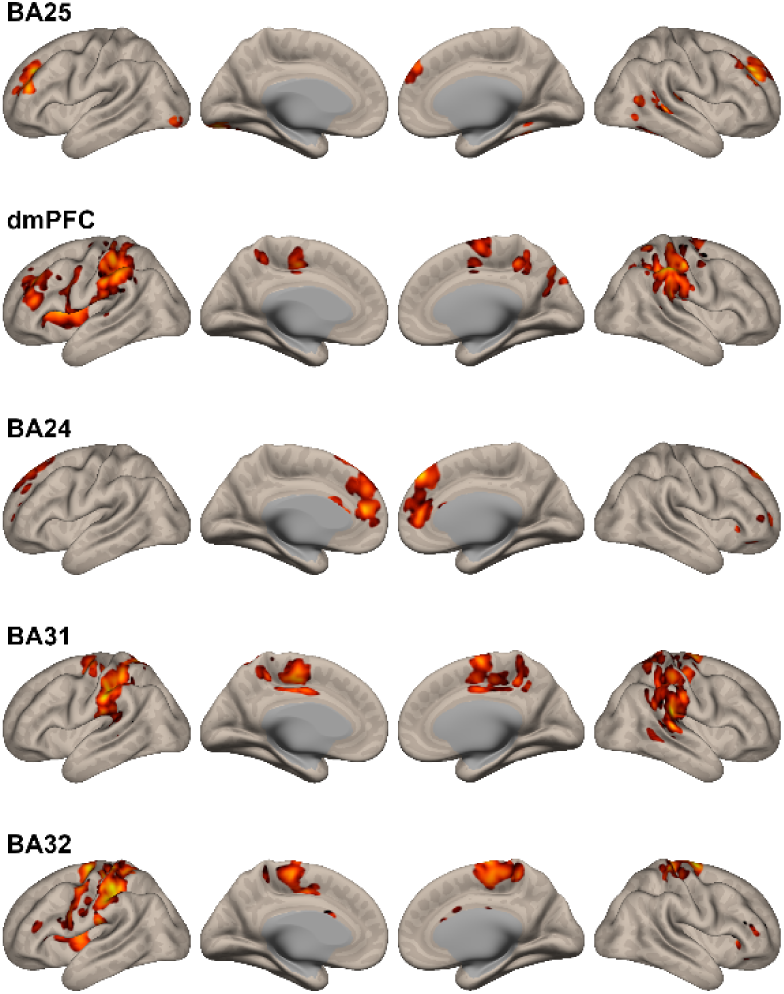
Changes in Functional Connectivity Observed After Placebo Exposure. A distinctly different pattern from N_2_O exposure was observed across all five seeds. Except for the cingulo-opercular seed (BA 24), functional connectivity changes observed following placebo exposure did *not* involve networks associated with mood regulation.

### Subgenual Cingulate Seed (BA25)

Following N_2_O exposure, we observed functional connectivity changes in the left subgenual cingulate seed -> voxel maps (TRD > CNTL; **Figure 3, row 1),** localized to bilateral medial prefrontal/paracingulate areas (BA 32), bilateral dorsolateral prefrontal cortices (BA 9), left lateral orbitofrontal cortex (BA 47), and anterior insular cortical regions. The TRD participants initially had higher baseline connectivity, which decreased after N_2_O exposure. In contrast, CNTL showed lower baseline connectivity that increased after N_2_O exposure (**Figure 5, top**).

**Figure 5.**
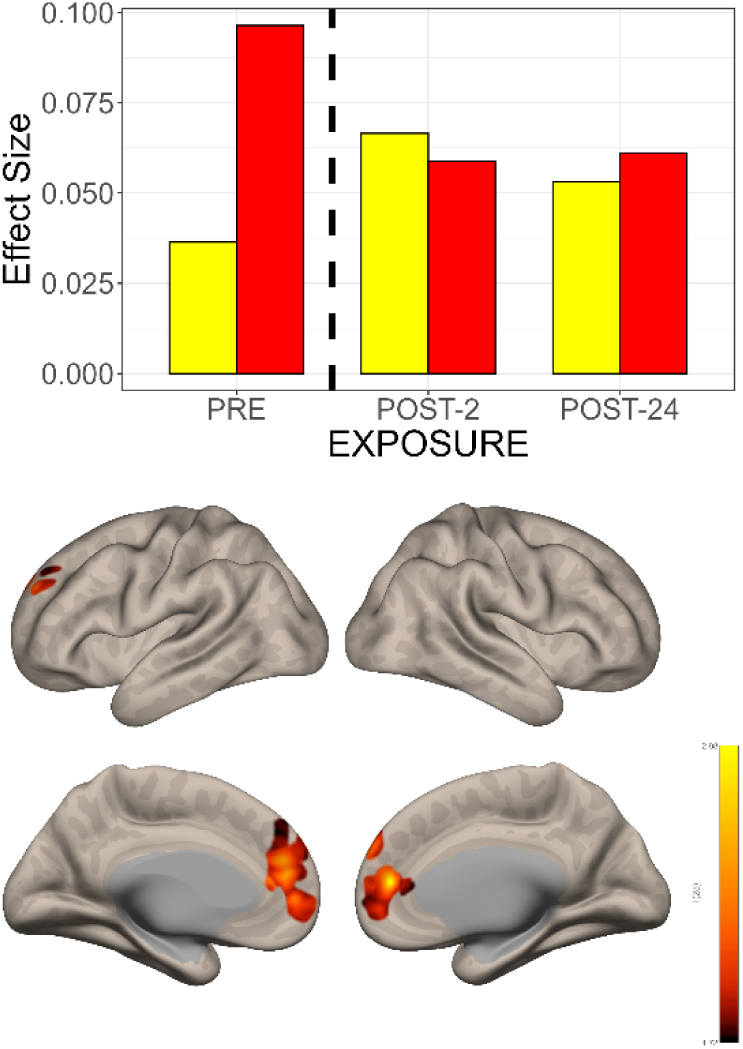
The Interaction Between Time and Group Shows Different Changes in Functional Connectivity in the Subgenual Cingulate Seed->Voxel Map. <u>TOP</u>: Effect size changes in functional connectivity between subgenual cingulate (BA 25) and a cluster in the dorsal paracingulate gyrus. Pre-exposure baseline connectivity was higher for the TRD patients (red bar) than the CNTL (yellow bar). Over the 2-24 hours following N_2_O exposure, the functional connectivity between BA25 and the dorsal paracingulate cluster *decreased*, approaching that of the CNTL. The opposite pattern was observed in the CNTL i.e., with time there was a gradual *increase* in functional connectivity. <u>BOTTOM:</u> Comparison of pre-versus post-exposure N_2_O effects on functional connectivity over time demonstrates changes in the bilateral medial prefrontal, anterior cingulate, paracingulate cortex and the dorsal paracingulate.

Left subgenual cingulate seed -> voxel map changes occurring following placebo exposure were quite different spatially (**Figure 4, Row 1**), and included bilateral cerebellar cortex, bilateral lingual gyrus (BA 19) and bilateral insula, and medial temporal regions including the hippocampus and parahippocampus. Of note, placebo produced *no functional connectivity changes in the dorsal paracingulate gyrus*, as observed following N_2_O exposure.

### Ventral Anterior Cingulate Seed (BA24)

Following N_2_O inhalation, functional connectivity changes were observed in the ventral anterior cingulate seed -> voxel maps (TRD > CNTL; **Figure 3, Row 3)** in the bilateral anterior frontal poles (BA10) and dorsal anterior cingulate (BA 32), as well as right paracingulate cortex (BA10). anterior frontal pole (BA10), and dorsal anterior cingulate (BA32).

### Dorsal Anterior Cingulate Seed (BA32)

Following N_2_O inhalation, functional connectivity changes were observed in the dorsal anterior cingulate seed -> voxel maps (TRD > CNTL; **Figure 3, Row 5),** in the right frontal pole (BA10), anterior cingulate and paracingulate gyrus (BA 24 and 10, respectively), middle frontal gyrus (BA 8), and precentral gyrus and left anterior insular cortex.

### Default Mode Seeds

#### Anterior - Ventromedial Prefrontal Cortex Seed (BA 10/14)

Following N_2_O inhalation, functional connectivity changes were observed in ventromedial PFC seed -> voxel maps (TRD > CNTL; **Figure 3, Row 2)** in the left hemisphere temporal pole (BA 38), middle temporal gyrus (BA 21), and orbitofrontal cortex (BA 47). Right hemisphere connectivity changes included the right parahippocampal gyrus (BA 36) and right hippocampus.

#### Posterior - Posterior Cingulate Seed (BA 23/31)

Following N_2_O inhalation, functional connectivity changes were observed in the posterior cingulate seed -> voxel maps (TRD > CNTL; **Figure 3, Row 4)** in the left precuneus, superior parietal lobule, post central gyrus, and lateral occipital cortex, as well as the right superior parietal lobule.

#### Common Spatial Effects

Following N_2_O exposure, all of the seed -> voxel maps overlapped in the dorsal paracingulate cortex (**Figure 3**). In contrast, following placebo exposure, none of the seeds were associated with change in this region (**Figure 4**).

#### Global Correlation

Pre-post N_2_O exposure global correlation (GCOR) of network functional connectivity changes (TRD > CNTL) demonstrated functional connectivity changes that were relatively symmetrical across hemispheres and included bilateral dorsal anterior cingulate (BA 32), frontal poles (BA10), left dorsolateral prefrontal cortex (BA9), right superior frontal gyrus (BA8) and orbitofrontal cortex (BA9) (**Figure 6, bottom**).

**Figure 6.**
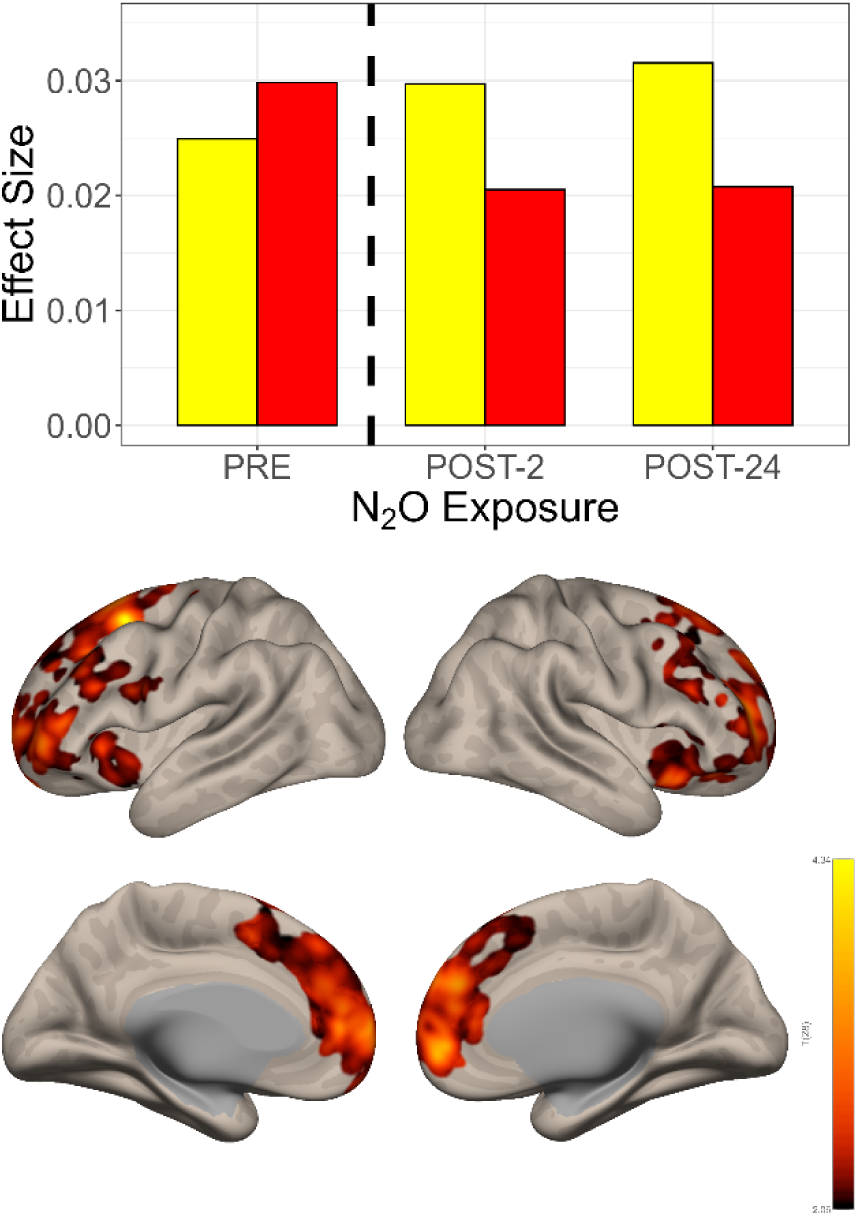
Global correlation changes (GCOR) associated with N_2_O exposure. <u>Top</u>: Bar graph of effect size changes in functional connectivity over time. Similar to the individual seed -> voxel maps, functional connectivity changes were in opposite directional following N_2_O exposure: CNTL participants (yellow bars) demonstrated *increased* functional connectivity and TRD participants (red bars) *decreased* functional connectivity. <u>Bottom</u>: GCOR changes associated with N_2_O exposure in the combined sample (CNTL + TRD) included bilateral dorsal anterior cingulate, frontal poles, left dorsolateral prefrontal cortex, and right superior frontal gyrus and orbitofrontal cortex.

A similar analysis was run using placebo exposure and we found no clusters of significant change (TRD > CNTL) in functional connectivity. Similar to the changes observed associated with N_2_O exposure in the seeded analyses, the post-N_2_O GCOR changes were in opposite directions: increases in connectivity over time were observed in the CNTL participants, whereas TRD participants demonstrated decreased connectivity (**Figure 6, top**).

#### Head Motion

Imaging data of the two cohorts were assessed for differences in head motion measures. Examination of group differences in inter-scan head motions measures did not reveal statistically significant group differences and measures were all lower in the TRD group. Mean inter-scan head motion was 0.011mm lower in the TRD group (95% CI −0.043 - 0.067). Maximum inter-scan head motion was −0.13 mm lower in the TRD group (95% CI −1.62 - 1.36). Maximum inter-scan global signal change was −1.60 units lower in the TRD group (95% CI - 10.12 - 13.33).

#### Behavioral outcomes

There was a non-significant greater reduction in depression symptoms (as measured by HAMD-17) in the participants receiving N_2_O over placebo. Similar to other nitrous oxide trials (1,2), significant placebo effects (**Figure 7, left panel**), as well as powerful carry-over effects (**Figure 7, right panel**), were observed despite a minimal one-month separation between inhalation sessions, i.e., those TRD participants randomized first to N_2_O were more likely to undergo the 2^nd^ inhalation (placebo) with a *lower* depression score than those starting with placebo (**Figure 7, right panel).** No N_2_O-placebo differences were observed in psychosis (BPRS) or dissociation (CADSS-28) measures. **Supplementary Table 4** summarizes changes in depression ratings.

**Figure 7.**
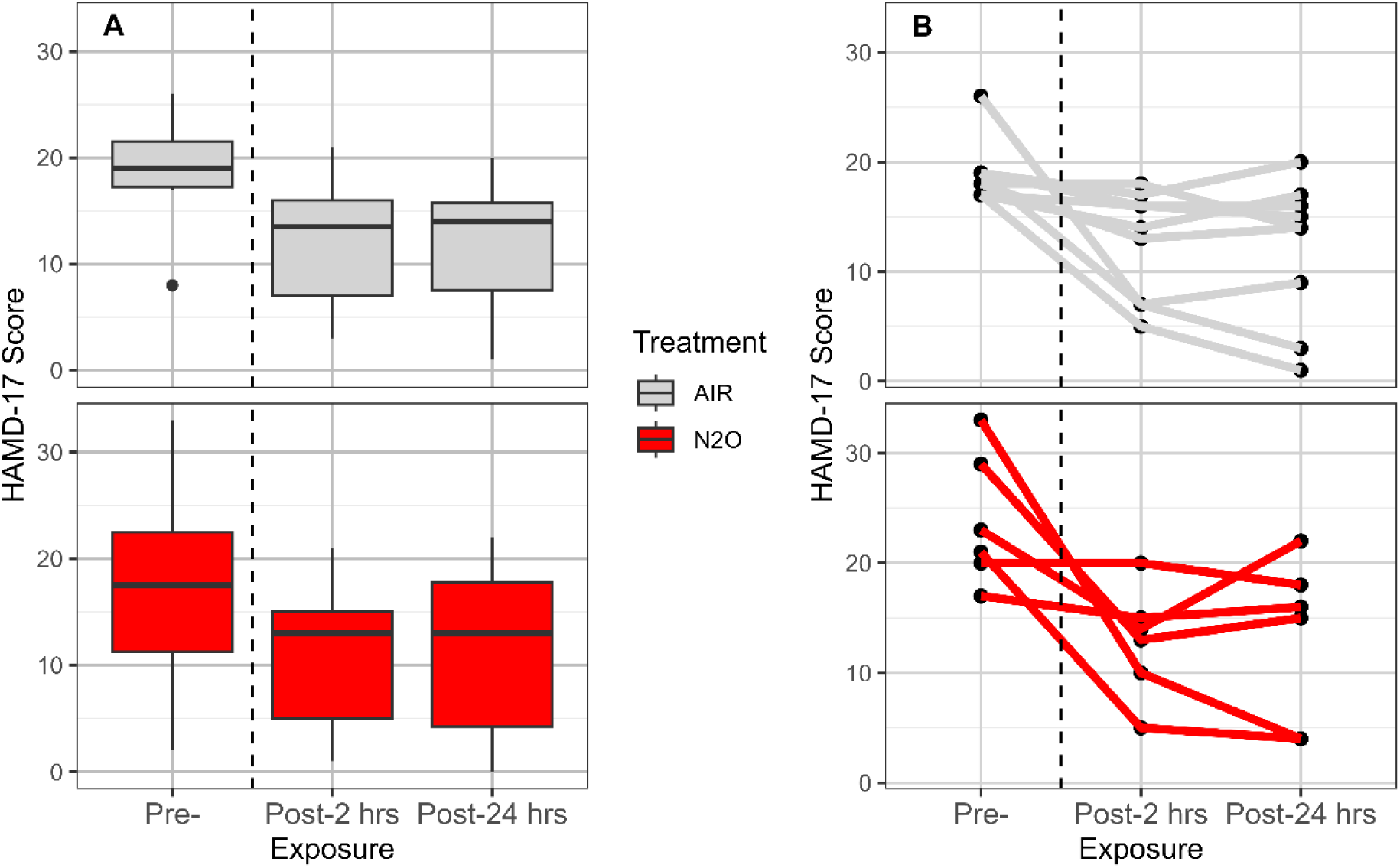
Depressive symptom changes pre-post N_2_O and placebo exposure. <u>Left Panel</u>. Both N_2_O and placebo exposure resulted in a reduction in depressive symptoms of TRD participants. Previous studies (1, 2) have demonstrated powerful placebo effects that decrease with time after exposure. <u>Right Panel</u>. In comparing the reduction in depressive symptoms of those participants who received either treatment *first*, those receiving N_2_O first demonstrated larger reductions in depressive symptoms.

#### Blinding Efficacy

The assay of the subjects’ certainty concerning the blind revealed that subjects did not correctly identify which exposure they were receiving. Fisher’s exact test was used to determine whether there was a significant association between gas mixture exposure type and the participants’ certainty rating. There was not a statistically significant association between the two variables (two-tailed p = 0.45).

## DISCUSSION

N_2_O inhalation, but not placebo, was associated with sustained functional connectivity changes in the dorsal paracingulate cortex (“dorsal nexus”), a region associated with mood regulation (18). A consistent pattern of depression-dependent functional connectivity changes was observed: N_2_O exposure *decreased* functional connectivity in TRD participants over time, whereas N_2_O exposure *increased* functional connectivity in CNTL participants. These depression-dependent temporal trends were observed in all seed -> voxel maps, as well as the global correlation (GCOR) analysis. Of particular note, the subgenual cingulate seed (RN) in the TRD cohort demonstrated considerably higher baseline connectivity (Figure 5**, top panel**) vis-à-vis the CNTL cohort. However, following N_2_O inhalation, this connectivity difference was reduced to align more with the CNTL cohort, suggesting that N_2_O may “normalize” functional connectivity in this circuit critical in MDD/TRD.

In general, N_2_O-associated TRD brain functional connectivity changes also exhibited a different spatial distribution than those associated with placebo. Although there were some functional connectivity changes observed with placebo, these changes *did not* involve medial prefrontal and anterior insular cortical regions affected by N_2_O exposure. This is critical, as these regions (encompassing the DMN and SN) are known to be central in MDD (12). Further, GCOR analyses of N_2_O-associated functional connectivity changes, which represent the areas of the entire brain undergoing the greatest amount of long-range connectivity change following N_2_O exposure, also demonstrated connectivity changes primarily affecting these medial-prefrontal regions (Figure 6). These GCOR connectivity changes were also *not* observed with placebo. In total, these findings demonstrate that inhaled N_2_O has differential effects on brain networks in TRD patients (vis-à-vis CNTL), more specifically, in several networks (DMN, SN, and RN) that are important in mood disorders. Further, post-inhalation functional connectivity changes were observed for the “dorsal nexus” region for all five seeds. Of note, the GCOR measure also revealed similar patterns of change in the dorsal paracingulate cortex. This finding further supports that the “dorsal nexus” region is a critical crossroads among the depression-associated networks (18).

Other groups have demonstrated sustained prefrontal cortex changes with N_2_O. Similar to this trial, Desmidt et al. (46) used functional MRI (fMRI) in a group of mildly resistant depressed female patients (single antidepressant trial failure) 2 hours after inhaled N_2_O. This group found that antidepressant responders had a significant reduction in connectivity of the subgenual anterior cingulate cortex (RN) with the posterior DMN (precuneus). Additionally, Desmidt et al. (46) employed a form of ultrasound (Tissue Pulsatility Imaging) for 10 minutes after N_2_O exposure and determined that N_2_O antidepressant responders, but not healthy controls or non-responders, had increased brain tissue pulsations, which correlated with cerebral blood flow. Critically, this trial did not include a placebo arm and, as noted below, placebo effects are substantial with N_2_O and ketamine; hence, a significant portion of the responders may not have actually responded to N_2_O. Kim et al. (5), using arterial spin labeling MRI with a sample of 25 bipolar I and II patients, found baseline (pre-treatment) low mean regional cerebral blood flow (rCBF) in anterior cingulate and ventral prefrontal cortex to be associated with 24-hour response to N_2_O.

Though too extensive to review comprehensively, ketamine TRD imaging studies have demonstrated findings consistent with those presented here, perhaps because both treatments engage NMDARs. These studies indicate that the anterior cingulate is critically involved in ketamine’s antidepressant effects (47). In a study with a very similar design to the current study, Alexander et al. (48) used rsfMRI analyses seeding the subgenual, perigenual, and dorsal anterior cingulate and found differential changes in connectivity with ketamine exposure in TRD versus non-depressed healthy controls. Similar to our N_2_O findings, this group reported depression-selective treatment effects: following ketamine exposure, connectivity of the subgenual cingulate-right hippocampus was *decreased* in TRD but *increased* in non-depressed controls. The authors emphasized the potential pivotal role of ACC regional differences in the action of ketamine, which may also be relevant for N_2_O.

Like previous N_2_O trials, when the two treatment arms were compared based on first exposure (to either N_2_O or placebo), N_2_O demonstrated greater antidepressant effects than placebo (Figure 7**, right panel**). Also, as observed in prior TRD N_2_O trials, there were significant placebo effects (1, 2). Of note, a recent meta-analysis demonstrated that ketamine has greater effects than placebo, even though 72% of the immediate and 66% of the antidepressant effects are potentially attributable to placebo (49). To date, existing data demonstrate that placebo effects observed with N_2_O are short-lived, resolving after one week or less (2). Such placebo effects are likely unavoidable for interventions that perturb the conscious state during administration. Persistence of antidepressant effects over weeks suggests that our findings are not strictly related to the placebo effect, however.

This study has several limitations. The small number of subjects, especially in the N_2_O arm, precluded the ability to analyze the interaction of N_2_O/placebo exposure connectivity changes and changes in depression scores. Consequently, our study was underpowered to assess for associations at an individual participant level. Despite this limitation, throughout the existing analyses of the combined group (TRD + CNTL), N_2_O-specific changes were observed in all of the MDD/TRD specific networks, which were not observed with placebo. Furthermore, we only assessed behavioral responses until 24 hours after N_2_O administration, but recently have shown that the strongest responses may last at least 2 weeks after administration (2). Additionally, given the powerful placebo effects observed in N_2_O trials (1, 2), and the observation that these placebo effects dissipate relatively rapidly, future placebo-controlled N_2_O depression studies should include behavioral measures at longer intervals (e.g., months after inhalation).

## CONCLUSION

Inhaled N_2_O induces sustained functional connectivity changes in depressed patients and healthy controls, but with opposite effect, showing a decrease in TRD patients, while an increase in healthy controls. These effects were observed in all depression-specific networks examined, manifesting overlap in the dorsal paracingulate cortex (“dorsal nexus”). Thus, spatiotemporal effects of N_2_O on long-range functional connectivity may underlie its anti-depressant action.

## MATERIALS AND METHODS

### Study Design

This prospective, placebo-controlled, double-blind crossover trial consisted of 2 inhalations over 6 neuroimaging sessions, distributed across 5 study visits (Figure 1).

A screening visit was used to verify eligibility and collect background information. The next visit consisted of depression assessment, a pre-exposure MRI scan session, a 60-minute gas exposure, and assessments 2 hours post-exposure with another MRI scan. The third visit, approximately 24 hours after gas exposure, included depression assessments and another MRI scan. The 2- and 24-hour post-exposure imaging times were chosen based on antidepressant response times observed in prior trials (1). Procedures were repeated 4-6 weeks later for the cross-over inhalation session, with imaging and assessments repeated on the same schedule as the first inhalation. Allocation (active treatment [50% N_2_O/oxygen] versus placebo [50% air/oxygen]) for the 2 arms was determined by a random number generator with block size of 2. The psychiatry team, who performed all diagnostic and depression scales, along with the TRD patients and CNTL participants, were unaware of treatment assignment. The anesthesia team was unblinded to the treatment and responsible for overseeing the randomization and conducting the inhalation sessions.

### Participants

Approval was obtained from the Washington University School of Medicine Human Research Protection Office and the study was prospectively registered on ClinicalTrials.gov (Identifier NCT02994433). Written informed consent was obtained from all study participants prior to any study activities and remuneration was provided for participants’ time and effort. Useable data was provided by 16 CNTL and 14 TRD patients; 13 TRD and 15 CNTL participants completed the full protocol.

Both TRD and CNTL groups were 18-65 years of age, right-handed, and had good command of the English language. Additionally, participants did not meet any of the following exclusion criteria: a primary DSM-IV Axis I diagnosis (other than major depressive disorder) per medical record review and Mini-international Neuropsychiatric Interview (MINI, 19); a severe personality disorder (as determined by clinical interview), or known primary neurological disorders or medical disorders affecting brain function, including dementia, stroke, encephalopathy, Parkinson’s disease, brain tumor, multiple sclerosis, or seizure disorder. Severe cardiopulmonary disease was also an exclusion. Additional exclusion criteria included: central nervous system active medication use (beyond existing antidepressant medications) or known disease affecting drug metabolism and excretion (e.g., renal or liver disease), as determined by the study investigator; ineligibility for MRI scans (e.g., a history of claustrophobia or MRI-incompatible metal implanted); recent (past 12 months) history of substance dependence or abuse, as determined by reported history and urine drug screen; or ability to become pregnant and not using effective contraception. Finally, participants could not have contraindications to N_2_O or study procedures, including pneumothorax, bowel obstruction, middle ear occlusion, elevated intracranial pressure, chronic cobalamin and/or folate deficiency treated with folic acid or vitamin B_12_, or current pregnancy or breastfeeding.

Additional inclusion criteria were group specific. TRD group inclusion required a _≥_17 score on the Hamilton Depression Rating Scale (HDRS) - 17 item (20) and failure to respond to _≥_3 lifetime adequate dose/duration antidepressant treatments with _≥_1 nonresponse in the current major depressive episode. CTNL participants had no history of depression as determined by report and structured clinical interview (MINI) and had to score _≤_7 on the HDRS-17. The CNTL group also had no current use of psychotropic medications, antidepressants, or prescription or nonprescription drugs/herbals intended to treat depression or anxiety.

### Inhalation sessions

Inhalation sessions were identical except for the composition of gas mixtures (50% N_2_O/oxygen or 50% air/oxygen). A previously effective antidepressant dose (50% N_2_O) and duration (1 hour) of N_2_O was employed (1). The gas mixture was administered via a standard anesthesia facemask through tubing connected to the anesthesia machine or via a facemask connected by a hose to an FDA-cleared Porter/Praxair MXR breathing circuit. A small sample connector line was used with the facemask to allow measurement of inhaled and exhaled gas concentrations, which were monitored and adjusted according to the protocol, gradually increasing N_2_O concentrations over the first 10 minutes of exposure. Total gas flow was maintained between 2-8 L/min. Participants were monitored according to American Society of Anesthesiologists standards, which included continuous 3-lead ECG, pulse oximetry, non-invasive blood pressure, and end-tidal carbon dioxide. After the inhalation, participants had vital signs and any potential adverse events monitored during a one-hour observation period.

### rs-fMRI Data Acquisition

Neuroimaging sessions utilized a Siemens 3T Prisma scanner with a 64-channel head coil. Whole-head T1-weighted structural images (1 mm^3^ voxels) were also acquired during the pre-inhalation session. For rs-fMRI, three 7.5-minute runs were acquired at each of the six imaging sessions, resulting in approximately 135 min of rs-fMRI data for each participant. For each run, Blood Oxygen Level Dependent (BOLD)-contrast multiband echo-planar imaging was acquired (2.4 mm^3^ voxels, echo time = 30 ms, repetition time = 0.8 seconds, 552 volumes/run). Participants were instructed to keep eyes open, remain relaxed and awake and focus on a fixation cross. FIRMM (Frame-wise Integrated Real-time MRI Monitoring; https://turingmedical.com/) software was used to monitor head motion. Additionally, four foam pads were utilized to minimize head movement during imaging.

### Image Analysis

#### Preprocessing

Preprocessing and subsequent analyses were performed using the CONN toolbox (21). Functional data were preprocessed using slice timing correction, realignment, outlier detection, Montreal Neurological Institute (MNI)-space direct normalization and smoothing. Temporal misalignment between different slices of the functional data (acquired in interleaved Siemens order) was corrected using the Statistical Parametric Mapping (SPM12) slice-timing correction procedure (22, 23) and sinc-temporal interpolation to resample each BOLD-contrast timeseries to the mid-volume slice time. Functional data were realigned using a least squares approach and a 6-parameter (rigid body) transformation without resampling (24). Head motion outlier scans were identified using the Artifact Reduction Tool (25). Time points with framewise displacement above 0.9 mm or global BOLD-contrast signal changes above 5 standard deviations (26, 27) were excluded. A mean BOLD-contrast image was computed for each subject by averaging all scans excluding outliers. Functional data were normalized into MNI space and resampled to 2 mm isotropic voxels using a direct normalization procedure (27–30) with the default IXI-549 tissue probability map template as a target. T1-weighted structural data were normalized into MNI space, segmented into grey matter, white matter, and CSF tissue classes, then resampled to 1 mm isotropic voxels (29, 30). Functional data were smoothed using spatial convolution with a Gaussian kernel of 8 mm full width half maximum (FWHM).

Next, functional data were denoised using a pipeline (31) that included the regression of potential confounding effects, leveraging the white matter timeseries (5 CompCor noise components), CSF timeseries (5 CompCor noise components), motion parameters and their first order derivatives (12 factors) (32), outlier scans (below 489 factors) (26), session and condition effects and their first order derivatives (6 factors), and linear trends (2 factors) within each functional run, followed by bandpass frequency filtering of the BOLD-contrast timeseries (33) between 0.008 Hz and 0.4 Hz. CompCor (34, 35) noise components within white matter and CSF were estimated by computing the average BOLD-contrast signal as well as the largest principal components orthogonal to the BOLD-contrast average, motion parameters, and outlier scans within each subject’s eroded segmentation masks. To minimize temporal edge effects, individual time series were weighted by a boxcar kernel convolved with a Hanning window.

#### First-level Modeling of Seed Effects

*A priori* cortical seed regions in medial limbic structures were used as registered with ClinicalTrials.gov (NCT02994433), Figure 2**, top panel.** These five seeds targeted: 1) the reward network (RN), composed of the subgenual cortex, ventral striatum and rostral anterior cingulate cortex (Brodmann Area 25, MNI seed coordinates −5,25,-10); 2) the anterior default mode network (DMN) (ventromedial prefrontal cortex, Brodman Areas 10/14, MNI seed coordinates −1,55,−3); 3) posterior DMN (posterior cingulate cortex and precuneus, Brodmann Areas 23/31, MNI seed coordinates −1,22,35); 3) the cingulo-opercular network (CON); 4) an executive control circuit including dorsal anterior cingulate cortex, dorsomedial prefrontal cortex, anterior insula, supramarginal gyrus, and pars marginalis of the cingulate gyrus (Brodmann Area 24, MNI seed coordinates −5,0,30); and 5) the salience network (SN), including portions of the anterior cingulate and anterior insular cortex (Brodmann Area 32, MNI seed coordinates −1, 22, 35).

Seed-based connectivity maps (SBC) characterized the patterns of functional connectivity using Fisher-transformed bivariate correlation coefficients from a weighted general linear model (36), modeling the associations in the seed->voxel BOLD-contrast signal timeseries, adjusted for physiological noise.

#### Group-level Modeling of Seed Effects

Second-level repeated measures models were performed using a General Linear Model (GLM) (37). Separate seed->voxel GLM contrasts were estimated, with first-level connectivity measures as dependent variables (one independent sample per subject for experimental condition (placebo or N_2_O) and group- or other subject-level identifiers as independent variables. Seed->voxel hypotheses were evaluated using parametric statistics. Inferences were performed at the level of individual clusters (groups of contiguous voxels). Cluster-level inferences were based on Gaussian Random Field theory (38, 39). Results were thresholded using a threshold-free cluster estimation (TFCE) p < 0.05 critical threshold (40).

#### Global Correlation Mapping

To elucidate other N_2_O-associated brain network exposure effects in addition to the seed-based results described above, voxel-wise global correlation maps (GCOR) were obtained. Our previous rs-fMRI analyses, limited to the effects of N_2_O in CNTL participants, had demonstrated persistent N_2_O exposure effects using this measure (10).

GCOR maps characterizing network centrality at each voxel were estimated as the average of all short- and long-range voxel-wise correlations among each voxel and the rest of the brain. Connections were computed from the matrix of bivariate correlation coefficients between the BOLD-contrast timeseries from each pair of voxels, estimated using a singular value decomposition of the Fischer z-score normalized BOLD-contrast signal (subject-level SVD) with 64 components separately for each subject and condition.

#### Behavioral Assessments and analyses

Standard scales were administered at each visit to measure the effects of the inhaled gases on behavioral and mood states, which included clinician-administered scales by one of the blinded psychiatry study team members. The Hamilton Depression Rating Scale-17 item (HDRS-17) served as the primary outcome measure. Secondary behavioral outcome measures included the Montgomery-Asberg Depression Rating Scale (MADRS, 41), the Clinical Administered Dissociative States Scale 28 item (CADSS-28, 42), the Brief Psychiatric Rating Scale (BPRS, 43), and self-report scales: Quick Inventory of Depressive Symptomology-Self Report (QIDS-SR, 44) and the Profile of Mood States 2nd Edition (POMS-2, 45). Additionally, adequacy of the blind was assessed by a self-administered 5-point Likert scale given after completion of inhalation sessions.

After each gas mixture exposure session, participants were asked to use a Likert Scale to rate the likelihood that they had received either mixture. Responses were: I strongly believe the treatment was nitrous oxide, I somewhat believe the treatment was nitrous oxide, I somewhat believe the treatment was placebo, or I strongly believe the treatment was placebo.

#### Statistical Analyses of Behavioral Measures

We fit a repeated-measures, linear mixed model (estimated using REML and the Nelder-Mead optimizer) to predict HAMD-17 Score with Treatment (N_2_O vs placebo), Session and Order (formula: HAMD-17 Score ∼ Treatment + Session + Order). The model included Participant ID as a random effect (formula: ∼1 | Participant ID). We obtained standardized parameters by fitting the model on a standardized version of the dataset. 95% Confidence Intervals (CIs) and p-values were computed using a Wald t-distribution approximation.

## Data Availability

All data associated with the study are present in the paper or the Supplementary Materials.

## Acknowledgments

We thank the Taylor Family Institute for Innovative Psychiatric Research at Washington University School of Medicine in St. Louis for their support. We also thank the clinical staff from Washington University and Barnes-Jewish Hospital. We thank the following colleagues for administering our inhalation sessions: Branden Yee, Sara Clarkson, James Fehr, Mohammad Helwani, James Hennessey, Mark Ingram, Helga Komen, Kristin Kraus, Michael Montana, Andreas Kokoefer, Jaime Brown-Shpigel, Christian Guay, Tracy Lanes, Sara Pitchford, and Marlette Williams.

## Author contributions

Conceptualization: CRC, BJP, TZ, CFZ, PN

Methodology: CRC, BJP, TZ, BMG, CFZ, PN

Clinical Trial and Data Acquisition: CRC, BJP, BMG, FB, VDL, LB, TN, WX, CLS, GE, PN

Imaging Data and Analysis: CRC, BJP, TZ,

Funding acquisition: CRC, BJP, CFZ, PN

Writing – original draft: CRC, BJP, TZ, CFZ, PN

Writing – review & editing: CRC, BJP, TZ, BMG, FB, VDL, LB, TN, WX, CLS, GE, SM, CFZ, PN

## Data and materials availability

All data associated with the study are present in the paper or the Supplementary Materials.

