## Supplemental_Materials for "Nitrous Oxide Alters Functional Connectivity in Medial Limbic Structures in Treatment-Resistant Major Depression"

### Additional Results

#### Figure 1S. Consort Diagram

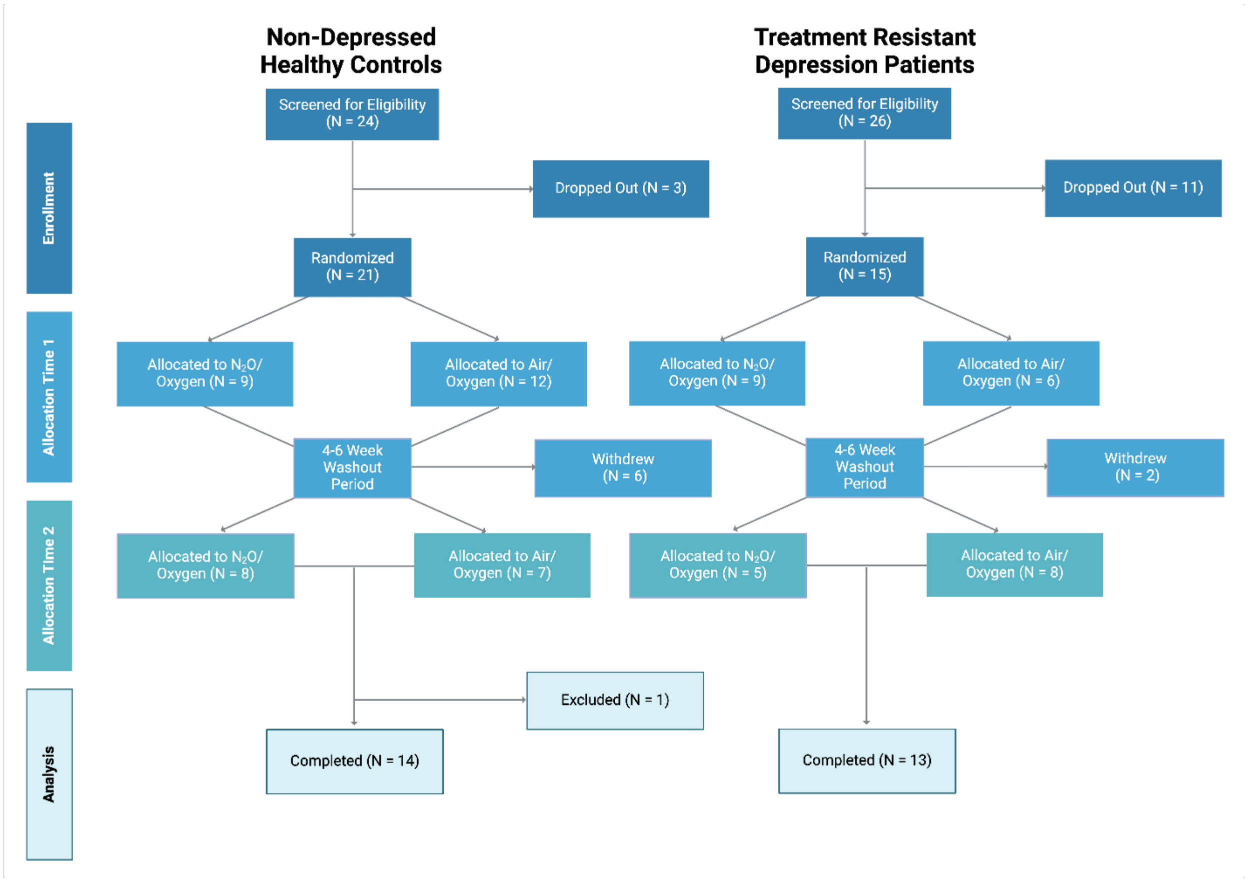


#### Participant Selection Process

Following eligibility screening, 24 CNTL and 26 TRD patients were consented via recruitment from an existing TRD database (gathered over the past 15 years at Washington University), existing psychiatric clinics at Washington University Department of Psychiatry, and through potential participant registries at Washington University School of Medicine and trial advertisement through the greater St. Louis community. Three CNTL and eleven TRD patients did not pass screening or voluntarily withdrew prior to randomization. Two TRD patients (one lost to follow-up, one terminated early due to Covid-19) and six CNTL (one lost to follow-up, one for mask discomfort, one determined to have pre-existing psychotic disorder, one could not attend sessions, one terminated early due to Covid-19, one due to nausea/dissociative experience) withdrew from the study following the first inhalation. Despite completing all inhalation and imaging sessions, 1 CNTL participant was excluded due to use of a different scanner system and imaging protocol.

**Table 1S. Treatment-Resistant Depressed Participant Demographics**

| Subject ID | Sex | Race | No. of trials, current episode | No. of failed trials, lifetime | Years reported with MDD |
|---|---|---|---|---|---|
| 1 | Male | White/Caucasian | 1 | 3 | 14 |
| 2 | Female | Asian | 1 | 3 | 7 |
| 3 | Male | White/Caucasian | 1 | 3 | 33 |
| 4 | Female | Black | 2 | 3 | 12 |
| 5 | Female | White/Caucasian | 1 | 9 | 38 |
| 6 | Male | White/Caucasian | 1 | 4 | 3 |
| 7 | Female | White/Caucasian | 2 | 19 | 39 |
| 8 | Female | White/Caucasian | 2 | 15 | 30 |
| 9 | Female | Asian | 1 | 3 | 3 |
| 10 | Female | White/Caucasian | 1 | 9 | 27 |
| 11 | Male | White/Caucasian | 1 | 4 | 19 |
| 12 | Female | White/Caucasian | 1 | 15 | 18 |
| 13 | Female | White/Caucasian | 2 | 14 | 36 |
| 14 | Female | White/Caucasian | 1 | 4 | 14 |

**Table 2S. Non-depressed Healthy Control Participant demographics**

| Subject ID | Sex | Race | Ethnicity |
|---|---|---|---|
| 1 | Female | White/Caucasian | Not Hispanic/Latino |
| 2 | Male | White/Caucasian | Not Hispanic/Latino |
| 3 | Male | Asian | Not Hispanic/Latino |
| 4 | Male | Asian | Not Hispanic/Latino |
| 5 | Female | Black | Not Hispanic/Latino |
| 6 | Female | White/Caucasian | Not Hispanic/Latino |
| 7 | Male | White/Caucasian | Hispanic/Latino |
| 8 | Female | Black | Not Hispanic/Latino |
| 9 | Male | Black | Not Hispanic/Latino |
| 10 | Female | White/Caucasian | Not Hispanic/Latino |
| 11 | Male | Black | Not Hispanic/Latino |
| 12 | Male | White/Caucasian | Not Hispanic/Latino |
| 13 | Female | White/Caucasian | Not Hispanic/Latino |
| 14 | Female | Black | Not Hispanic/Latino |
| 15 | Female | Black | Not Hispanic/Latino |
| 16 | Female | White/Caucasian | Not Hispanic/Latino |

**Table 3S. Differences in Network Connectivity in TRD and Healthy Controls**

| Seed and Associated Network | Seed Coordinates (MNI) | Cluster Size (# voxels) Cluster Peak Coordinates (MNI) |
|---|---|---|
| BA 25<br>Subgenual Cingulate Cortex<br>Reward Network seed<br>(Figure 4; Row 1) | -5,25,-10 | 664; L DLPFC (BA 9); -16,+54,+30<br>299; L Paracingulate Gyrus (BA 32);<br>-8,+48,+10<br>282; L Orbitolfrontal Cortex; (BA 47);<br>-38,+26,-8<br>274; R Paracingulate Gyrus (BA 32); +8,+50,+8<br>252; R DLPFC (BA 9); +6,+56,+10 |
| BA 10/14<br>Ventromedial Prefrontal Cortex<br>Anterior Default Mode Network seed<br>(Figure 4; Row 2) | -1,55,-3 | 452; L temporal pole (BA 38); -32,10,-30<br>312; L middle temporal gyrus, posterior division (BA 21); -56,-26,-6<br>186; R parahippocampal gyrus, posterior division (BA 36); 24,-28,-18<br>168; R hippocampus; 26,-20,-16<br>167; L frontal orbital cortex (BA 47); -28,22,<br>-22 |
| BA 24<br>Ventral Anterior Cingulate Cortex<br>Executive Control Network seed<br>(Figure 4, Row 3) | -5,0,30 | 436; R paracingulate cortex/anterior PFC (BA 10); 6,48,10<br>327; R anterior frontal pole (BA 10); 20,58,4<br>312; R dorsal ACC (BA32); 0,38,10<br>298; L dorsal ACC (BA 32); -6,46,14<br>264; L anterior frontal pole (BA 10); -18,58,-2 |
| BA 23/31<br>Posterior Cingulate Cortex<br>Posterior Default Mode Network seed<br>(Figure 4; Row 4) | -1,-61,38 | 684; L Precuneous Cortex (BA 31); -4,-54,48<br>590; R superior parietal lobule (BA 7); 28,-48,58<br>390; L superior pariental lobule (BA 7); -28,-46,58<br>259; L postcentral gyrus (BA 40); -42,-30,42<br>186; L lateral occipital cortex (BA 7); -22,-68,40 |
| BA 32 | -1,22,35 | 1868; R frontal pole (BA 10); 28,48,18.<br>1311; R middle frontal gyrus (BA 8); 36,16,46 |

| | | |
|---|---|---|
| Dorsal Anterior Cingulate Cortex<br><br>Salience Network seed<br><br>(Figure 4, Row 5) | | 622; R anterior cingulate gyrus (BA 24); 2,34,14<br><br>506; R precentral gyrus; 40,4,48<br><br>278; R paracingulate gyrus (BA 10); 12,42,16 |

**Supplementary Table 4S** Behavioral Assessments Following $N_2O$/Placebo Inhalation

| | Nitrous Oxide Timepoints | | | Placebo Timepoints | | |
|---|---|---|---|---|---|---|
| | 0 (baseline) | 2 hour post | 24 hour post | 0 (baseline) | 2 hour post | 24 hour post |
| **n** | | | | | | |
| controls | 16 | 16 | 16 | 17 | 17 | 17 |
| TRD | 14 | 14 | 14 | 14 | 14 | 14 |
| **Treatment = N2O (%)** | | | | | | |
| controls | 16 (100.0) | 16 (100.0) | 16 (100.0) | 0 (0.0) | 0 (0.0) | 0 (0.0) |
| TRD | 14 (100.0) | 14 (100.0) | 14 (100.0) | 0 (0.0) | 0 (0.0) | 0 (0.0) |
| **Sex = Female (%)** | | | | | | |
| controls | 7 (43.8) | 7 (43.8) | 7 (43.8) | 7 (41.2) | 7 (41.2) | 7 (41.2) |
| TRD | 4 (28.6) | 4 (28.6) | 4 (28.6) | 4 (28.6) | 4 (28.6) | 4 (28.6) |
| **MADRS (mean, SD)** | | | | | | |
| controls | 0.21 (0.58) | 0.29 (0.47) | 0.43 (1.09) | 1.24 (2.36) | 1.47 (2.58) | 0.88 (2.55) |
| TRD | 27.64 (15.18) | 16.21 (12.39) | 18.00 (13.36) | 30.21 (7.90) | 17.93 (10.62) | 16.29 (10.31) |
| **HAMD-17 (mean, SD)** | | | | | | |
| controls | 0.43 (0.76) | 0.21 (0.58) | 0.71 (1.33) | 1.24 (1.99) | 1.71 (2.39) | 1.12 (2.39) |
| TRD | 17.14 (8.87) | 11.23 (6.67) | 11.64 (7.35) | 19.07 (4.25) | 11.93 (5.77) | 11.71 (5.89) |
| **QIDS (mean, SD)** | | | | | | |
| controls | 2.36 (2.62) | 1.71 (1.54) | 1.50 (1.51) | 2.47 (1.50) | 1.94 (1.20) | 1.65 (1.32) |
| TRD | 13.43 (6.32) | 8.93 (6.16) | 9.93 (5.97) | 14.64 (4.22) | 9.64 (5.46) | 9.29 (5.88) |
| **CADSS (mean, SD)** | | | | | | |
| controls | 0.14 (0.36) | 0.07 (0.27) | 0.00 (0.00) | 0.29 (1.21) | 0.76 (2.91) | 0.76 (2.66) |
| TRD | 1.91 (4.30) | 1.00 (3.46) | 0.93 (2.64) | 0.93 (2.92) | 1.38 (3.48) | 0.50 (1.87) |
| **POMS (mean, SD)** | | | | | | |
| controls | 30.79 (9.82) | 28.69 (10.61) | 27.43 (9.98) | 31.80 (8.28) | 27.50 (7.70) | 26.75 (7.09) |
| TRD | 55.00 (23.61) | 34.00 (18.00) | 37.64 (18.25) | 55.38 (18.50) | 35.15 (16.00) | 36.15 (15.77) |
| **BPRS (mean, SD)** | | | | | | |
| controls | 24.21 (0.58) | 24.14 (0.53) | 24.29 (0.73) | 25.06 (2.28) | 24.94 (2.01) | 25.12 (2.12) |
| TRD | 37.08 (8.61) | 31.14 (6.97) | 32.38 (7.16) | 38.29 (5.37) | 31.14 (4.83) | 31.93 (5.47) |
